# Projecting the potential impact of an Omicron XBB.1.5 wave in Shanghai, China

**DOI:** 10.1101/2023.05.10.23289761

**Authors:** Hengcong Liu, Xiangyanyu Xu, Xiaowei Deng, Zexin Hu, Ruijia Sun, Junyi Zou, Jiayi Dong, Qianhui Wu, Xinhua Chen, Lan Yi, Jun Cai, Juanjuan Zhang, Marco Ajelli, Hongjie Yu

**Author notes:** Corresponding authors: Hongjie Yu. These authors are joint senior authors contributed equally to this work (Marco Ajelli, Hongjie Yu).

## Abstract

China experienced a major nationwide wave of SARS-CoV-2 infections in December 2022, immediately after lifting strict interventions, despite the majority of the population having already received inactivated COVID-19 vaccines. Due to the rapid waning of protection and the emergence of Omicron XBB.1.5, the risk of another COVID-19 wave remains high. It is still unclear whether the health care system will be able to manage the demand during this potential XBB.1.5 wave and if the number of associated deaths can be reduced to a level comparable to that of seasonal influenza. Thus, we developed a mathematical model of XBB.1.5 transmission using Shanghai as a case study. We found that a potential XBB.1.5 wave is less likely to overwhelm the health care system and would result in a death toll comparable to that of seasonal influenza, albeit still larger, especially among elderly individuals. Our analyses show that a combination of vaccines and antiviral drugs can effectively mitigate an XBB.1.5 epidemic, with a projected number of deaths of 2.08 per 10,000 individuals.

This figure corresponds to a 70–80% decrease compared to the previous Omicron wave and is comparable to the level of seasonal influenza. The peak prevalence of hospital admissions and ICU admissions are projected at 28.89 and 2.28 per 10,000 individuals, respectively, suggesting the need for a moderate increase in the capacity of the health care system. Our findings emphasize the importance of improving vaccination coverage, particularly among the older population, and the use of antiviral treatments.

## Background

China experienced a nationwide wave of COVID-19 dominated by the SARS-CoV-2 Omicron BA.5/BF.7 variants from December 2022 to January 2023, following the relaxation of stringent COVID-19 restrictions, known as the “zero-COVID” policy [1–3]. During this two-month period, an estimated 80–90% of the Chinese population was infected [4, 5]. This was in marked contrast to other parts of the world, where immunity was built through repeated vaccination and multiple waves of infection over more than two years [6]. As of April 27, 2023, 3.49 billion doses of the COVID-19 vaccine had been administered in mainland China, with 74% of adults (population over 18) having received their first booster [1]. Among the older population (population over 60), 73% had received their first booster [1]. However, recent studies have shown that protection against Omicron infection wanes within months after the previous vaccination or infection [7-10]. For example, Singapore experienced a 2-month XBB.1.5 wave in September 2022 following a BA.5 wave in May 2022 [11-13]. Therefore, even with high levels of population immunity in early 2023, China is still at risk of experiencing a major Omicron XBB.1.5 wave, which is particularly alarming given that the Chinese health care system was overwhelmed by the 2023 nationwide BA.5/BF.7 wave.

Since the BA.5/BF.7 wave, China has strengthened its health care capacity [14], and antiviral drugs such as Molnupiravir and Paxlovid and more than 10 domestically made products (e.g., RAY1216, XIANNUOXIN, and VV116) have either been granted official or conditional market approval or have been authorized to treat symptomatic patients [15-18]. Nevertheless, it is still uncertain if the strengthened health care system would be capable of managing the disease burden of a potential XBB.1.5 wave and if it could reduce mortality to a level similar to that of seasonal influenza. To address these questions, we used Shanghai as an example, developed a mathematical model of Omicron XBB.1.5 transmission and explored a set of mitigation strategies that combined the use of vaccines, antiviral drugs, and nonpharmaceutical interventions (NPIs).

## Methods

### Transmission model

We developed an age-structured stochastic model (Supplementary method) to simulate the transmission of SARS-CoV-2 Omicron XBB.1.5. The model focuses on the Shanghai population and accounts for heterogeneous contact patterns [19]. In the model, the population is divided into four epidemiological categories: susceptible, latent, infectious, and temporarily protected (removed) individuals, stratified by 9 age groups (0-2, 3-17, 18-29, 30-39, 40-49, 50-59, 60-69, 70-79 and 80+). Briefly, upon infection, susceptible individuals enter a latent compartment before becoming infectious. Infectious individuals can potentially infect susceptible individuals, and the generation time was set at 5.9 days based on a pooled estimate (Supplementary Figure 1) [20-22].

A total of 1,380 imported XBB.1.5 infections within 10 days were considered, and 6-month forward projections were provided. The average daily number of imported XBB.1.5 infections was estimated based on the average daily imported COVID-19 cases in November 2022 and scaled by the ratio of airport passenger traffic between November 2022 (0.14 million) and February 2023 (0.55 million) [23-25]. We conducted sensitivity analyses considering different seeding dates (July 1, August 1, and September 1) and the number of imported infections (500 and 2000 within 10 days).

Shanghai experienced a COVID-19 wave dominated by Omicron BA.5 in the winter of 2022. Approximately 80-90% of the population was infected within 2 months [4]. Therefore, a homogeneous hybrid immunity of 85% generated by inactivated vaccine and BA.5 infection was considered. The remaining 15% of the population was assumed to be fully susceptible to infection. Immunity was assumed to wane over time, beginning in February 2023, with different waning rates depending on the clinical endpoints. We used an average duration of protection against infection of 6 months following an exponential waning rate [10]. Hybrid immunity provides more significant and long-lasting protection against hospitalization and severe disease [9, 10]. As such, we assumed protection against hospitalization and severe disease to be 95% 12 months after the last infection or vaccination. Sensitivity analyses on the waning rates were also conducted (Supplementary Table 1).

The effective reproduction number was set at 3.4, similar to the values estimated for the Omicron BA.2 and BA.5 outbreaks in Shanghai [26, 27]. Given that the interplay between transmissibility and immune evasive characteristics of the virus and the waning of immunity in the population are hard to project, we considered two additional scenarios with effective reproductive numbers set at 3.0 and 3.7 as sensitivity analyses.

### Disease burden

Intrinsic age-specific infection-hospitalization risk (IHR), infection-ICU risk (IUR), and infection-fatality risk (IFR) were used to estimate the disease burden (i.e., number of hospital admissions, ICU admissions, and deaths) given the number of infections projected by the transmission model. The overall IHR and IFR of BA.5 were estimated to be approximately 4.1% and 0.5%, respectively, in China [28]. A similar intrinsic clinical severity of BA.1 (i.e., IHR=3.5%, IFR=0.6%) was reported by *Perez-Guzman et al* [29]. According to the World Health Organization, XBB.1.5 does not show signs of change or increase in clinical severity [30]. Therefore, we used the rates estimated for BA.5 (Supplementary Table 2) to model the burden of XBB.1.5. We considered the time periods between infection and hospital admission and from infection to death to follow two gamma distributions with means of 6.7 and 22.3 days, respectively [31-33]. The length of hospital stay (regardless of whether the patient required ICU admission) was set at 8 days [34]. The number of hospital beds and ICU beds designated for COVID-19 patients was 19.6 and 2.0 per 10,000, respectively (which corresponds to approximately 30% of total hospital capacity and 15% of total ICU capacity) [28, 35, 36].

### Pharmaceutical interventions

#### Vaccination

We assumed that the booster immunization campaign would continue, with 70% of adults who had hybrid immunity receiving a heterologous bivalent booster shot 3 months after recovering from a previous infection [1, 37-39]. We set the daily booster vaccination rate at 0.5%, which is equivalent to 30% of the maximum daily vaccination rate during the rapid rollout of mass vaccination in 2021 [40]. We assumed that the bivalent booster would provide a slightly higher level of protection against clinical endpoints than hybrid immunity (Supplementary Table 1). Furthermore, we examined alternative scenarios where 30% and 90% of eligible adults would receive a bivalent vaccine as a booster shot 3 months after the previous vaccination or recovery from a previous infection (Supplementary Table 1). In addition, we conducted a sensitivity analysis in which 70% of eligible adults received a monovalent booster 3 months after their previous vaccination or infection (Supplementary Table 1).

#### Antiviral drugs

We assumed that 10% of adult patients would receive antiviral treatment using nirmatrelvir/ritonavir. This is in line with what has been observed in Hong Kong since mid-March 2022, when nearly 60% of eligible symptomatic patients received antiviral treatment [28]. We examined two additional scenarios to evaluate the effect of antiviral drugs: (1) no patients would receive antiviral treatment, and (2) 20% of adult patients would receive antiviral treatment. For individuals who received combined nirmatrelvir/ritonavir antiviral treatment, their risks of hospitalization and death were reduced by 24% and 66%, respectively [41].

#### Nonpharmaceutical interventions

We modeled two types of NPIs: 10% home isolation and L1 PHSMs (level 1 public health and social measures). Home isolation entailed isolating infectious individuals at their place of residence and lowering their transmissibility by an estimated 70% [28]. L1 PHSMs included measures such as voluntary universal face masking and improved hand hygiene, and these measures were found to decrease the reproduction number by 15% [28]. The home isolation probability factored into the test sensitivity the probability of being tested if infected and the probability of complying with the home isolation policy. We also examined three alternative scenarios: (1) no isolation, (2) 20% of all infected individuals isolated at home after a positive rapid antigen test result, and (3) no L1 PHSMs implemented.

#### Statistical analysis

For each scenario, 100 stochastic model realizations were performed. We estimated median and 95% confidence intervals as the 2.5th and 97.5th percentiles of the distribution of the analyzed quantity derived from the 100 stochastic model realizations.

## Results

Our reference scenario considers 70% booster vaccination coverage, 10% antiviral drugs, 10% isolation and L1 PHSMs. Our projections for this scenario indicate that the emergence of Omicron XBB.1.5 would trigger a major two-month wave in Shanghai, resulting in 132,564 hospital admissions (51.23 per 10,000), 10,967 ICU admissions (4.24 per 10,000), and 4,387 deaths (1.70 per 10,000 compared to 1.3 deaths per 10,000 estimated for seasonal influenza [42]) (Fig. 1A-I). The burden of the outbreak is projected to be heaviest on the population over 60 years old, accounting for nearly 40% of hospital admissions (Fig. 1B) and 90% of ICU admissions and deaths (Fig. 1E, H). Specifically, this age group is projected to experience 6.79 deaths per 10,000 compared to 3.8 deaths per 10,000 estimated for seasonal influenza [42]. Our projections also suggest that the epidemic peak would require 24.2 hospital beds per 10,000 residents (Fig. 1A), exceeding the maximum daily hospital capacity dedicated to COVID-19 (19.6 per 10,000). Similarly, 1.7 ICU beds per 10,000 residents would be required at the epidemic peak (Fig. 1D), which falls under the maximum daily ICU capacity designated for COVID-19 (2.0 per 10,000). The period of hospital bed shortages is estimated to last for approximately 8 days (Fig. 1A). Consistent results were found for all our sensitivity analyses considering different seeding dates, numbers of imported infections, values of the effective reproduction number, waning rates of immunity, and types of booster vaccines (Supplementary Figure 2-6).

**Figure 1.**
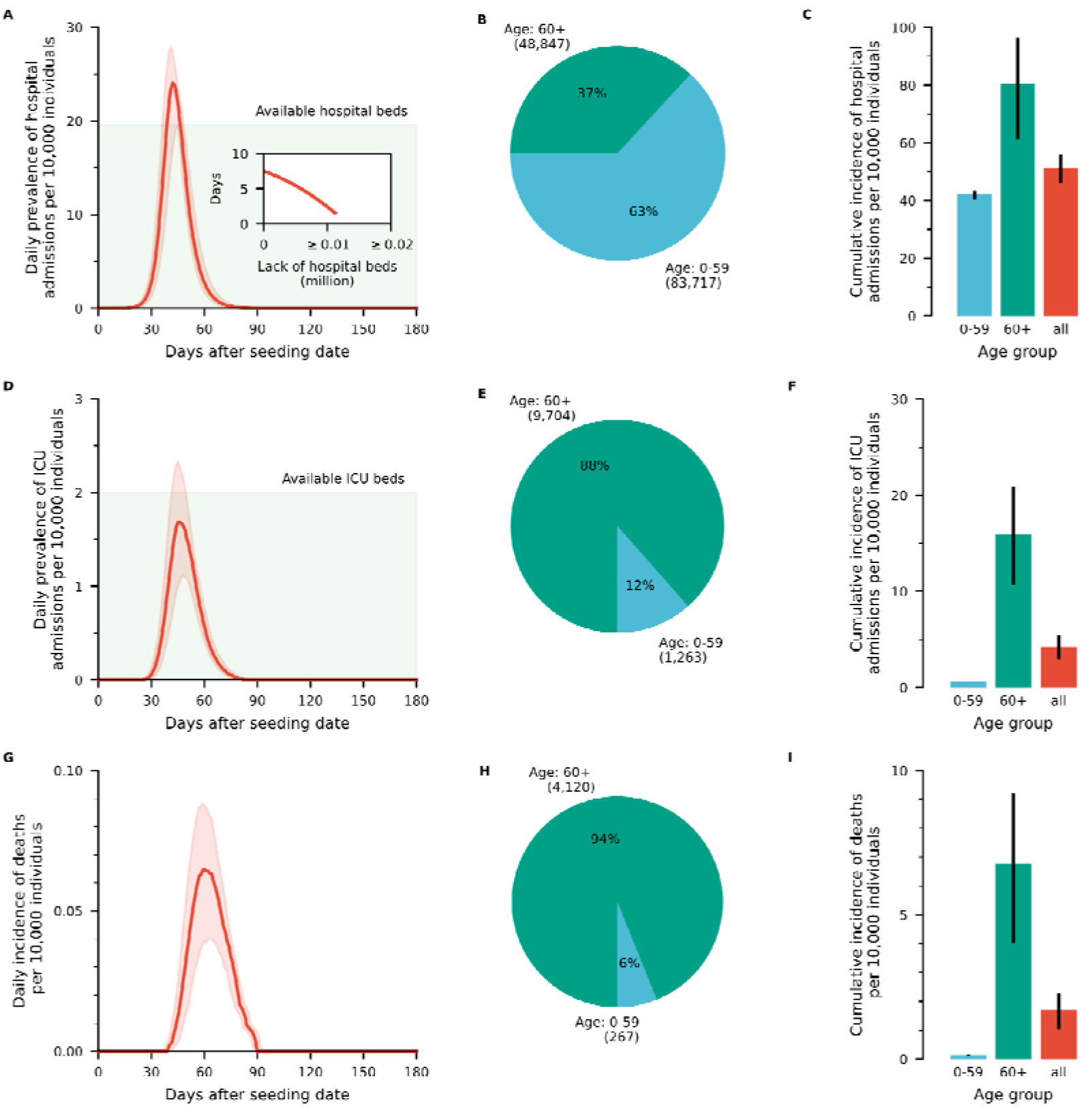
Projected COVID-19 burden for the reference scenario in which 70% booster vaccine coverage, 10% antiviral drugs, 10% home isolation, and L1 PHSMs are implemented. **A** Daily prevalence of hospital admissions per 10,000 individuals. The shaded area corresponds to the maximum hospital bed capacity. The inset shows the number of days of hospital bed shortage as a function of the number of missing beds. **B** Distribution of hospital admissions by age group. **C** Cumulative number of hospital admissions by age group per 10,000 individuals in that age group. **D** Daily prevalence of ICU admissions per 10,000 individuals. The shaded area corresponds to the maximum ICU capacity. **E** Distribution of ICU admissions by age group. **F** Cumulative number of ICU admissions by age group per 10,000 individuals in that age group. **G** Daily incidence of deaths per 10,000 individuals. **H** Distribution of deaths by age group. **I** Cumulative number of deaths by age group per 10,000 individuals in that age group. Data are presented as the median and 95% CIs of 100 stochastic model realizations.

We then investigated the impact of varying the intensity of each intervention separately (Fig. 2 and Supplementary Figure 7). This analysis underscores the importance of maintaining certain levels of NPIs. For instance, if L1 PHMS were to be lifted, the number of ICU admissions and deaths are projected to increase by 23% and 30%, respectively. Compared to the reference scenario, we found that doubling the isolation probability (from 10% to 20%), doubling the probability of receiving antiviral treatment (from 10% to 20%), or increasing vaccination coverage from 70% to 90% would reduce the projected number of ICU admissions and deaths by 10-20%.

**Figure 2.**
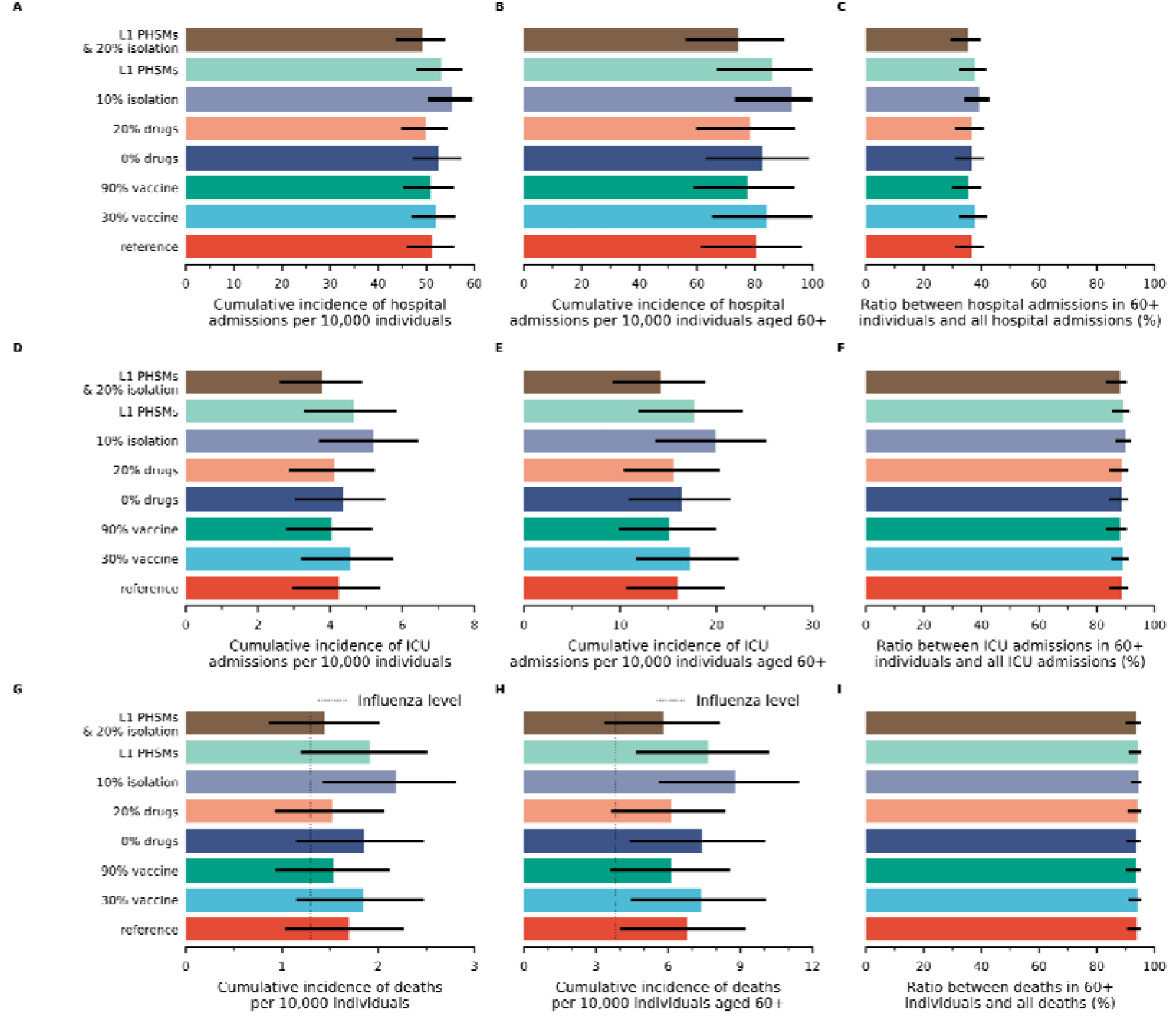
Projected impact on COVID-19 burden: Varying the intensity of each intervention separately. **A** Cumulative incidence of hospital admissions per 10,000 individuals. **B** Cumulative incidence of hospital admissions in 60+ individuals per 10,000 individuals aged 60+ years. **C** Ratio between hospital admissions in 60+ individuals and all hospital admissions. **D-F** Same as A-C, but for the ICU. **G-I** Same as A-C, but for death. The dotted line in Panels G and H represents the corresponding influenza-related excess mortality. *Reference* represents the reference scenario considering that 70% booster vaccine coverage, 10% antiviral drugs, 10% home isolation, and L1 PHSMs have been implemented. The other scenarios are obtained by changing a single parameter at a time compared to the reference scenario. Data are presented as the median and 95% CIs of 100 stochastic model realizations.

Next, we evaluated the synergetic effect of multiple interventions (Fig. 3 and Supplementary Table 4). A combination of high booster coverage (90% coverage) with antiviral treatment (20% receive antiviral treatment) would be enough to reduce the number of deaths to levels comparable to those of seasonal influenza (Fig. 3A-B). The additional use of the moderate level of NPIs (20% home isolation and L1 PHSMs) would be needed to lower the number of deaths to a level close to seasonal influenza, albeit still slightly higher for 60+ individuals (Fig. 3A-B). This combination would also considerably lower the demand for hospital beds and ICU beds (Fig. 3C-D). Although additional hospital beds may be needed to meet the demand (Fig. 3C), this combined strategy is projected to prevent ICU from being overwhelmed (Fig. 3D).

**Figure 3.**
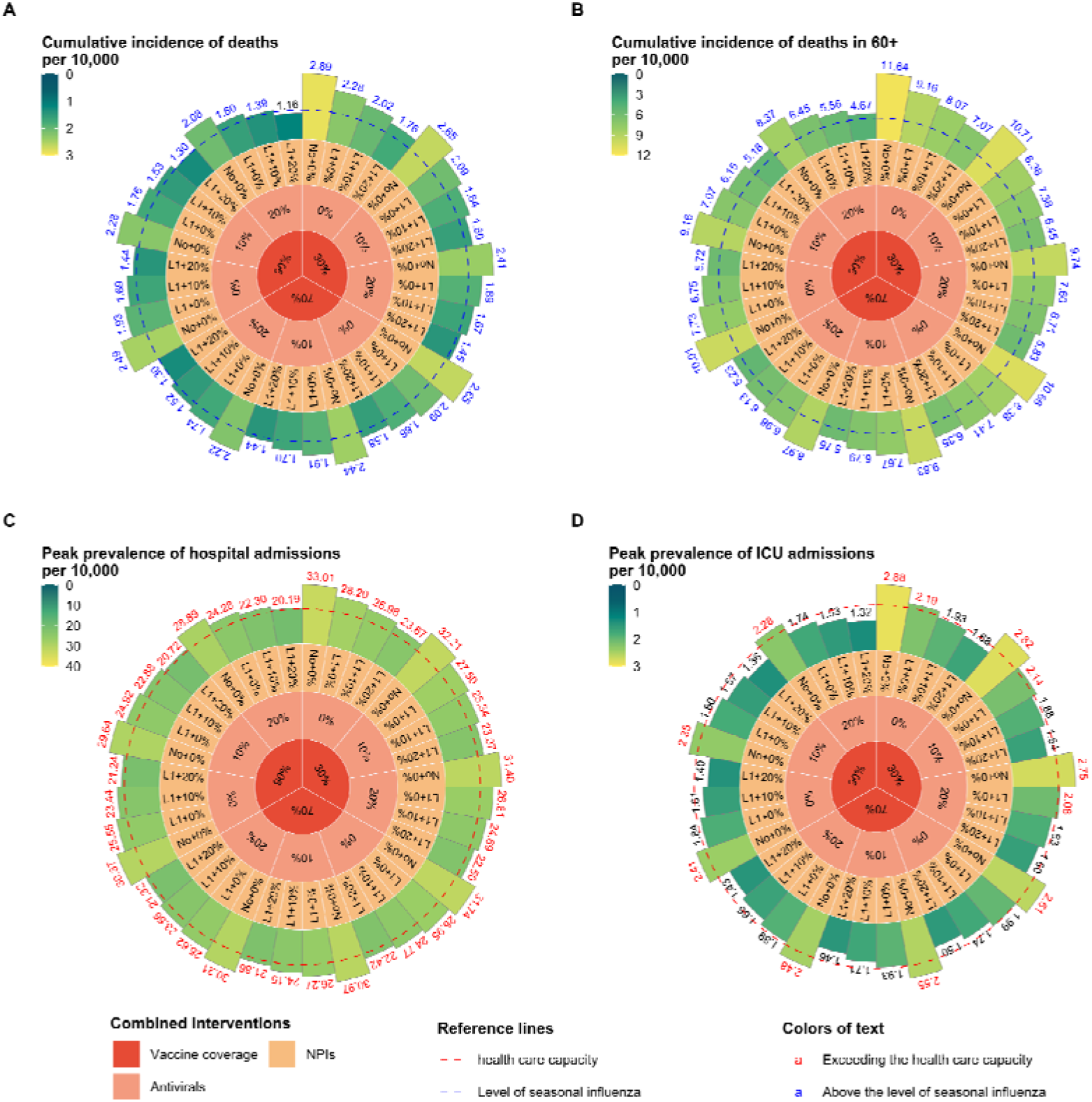
Projected health care demand and the number of deaths when adopting combined interventions. **A** Cumulative incidence of deaths per 10,000 individuals. **B** Same as A, but for individuals aged 60+. **C** Peak prevalence of hospital admissions per 10,000 individuals. **D** Same as C, but for ICU. No+0%: no NPIs. L1+0%: L1 PHSMs. L1+10%: L1 PHSMs and 10% probability of home isolation. L1+20%: L1 PHSMs and 20% probability of home isolation. Data are presented as the median and 95% CIs of 100 stochastic model realizations.

## Discussion

In this study, we developed a stochastic compartmental model of SARS-CoV-2 transmission to project the potential impact of an Omicron XBB.1.5 epidemic wave in China using Shanghai as a case study. In the reference scenario, our projections highlight the potential for a major 2-month epidemic wave, which would possibly require a moderate expansion of the health care capacity to meet the demand for hospital beds and ICU beds. Specifically, our study estimates a peak need of 24.2 hospital beds per 10,000 individuals (compared to a health care capacity of 19.6 per 10,000) and 1.7 ICU beds per 10,000 individuals (compared to a health care capacity of 2.0 per 10,000). Our analysis suggests that there would be 1.70 deaths per 10,000 individuals and 6.79 deaths per 10,000 individuals in the population over 60 years old, with more than 90% of deaths occurring in this age group. These figures represent a 1.3- and 1.8-fold increase compared to the annual influenza-related excess deaths reported between the 2010-11 and 2014-15 seasons, which were estimated at 1.3 and 3.8 deaths per 10,000, respectively [42].

The synergetic effect of vaccination (90% vaccination coverage) and antiviral treatment (20% receive antiviral treatment) could effectively mitigate an XBB.1.5 epidemic, reducing the number of deaths to a level comparable to that of seasonal influenza (2.08 per 10,000 residents and 8.37 per 10,000 individuals aged 60+). This combined strategy would also require a moderate expansion of the health care capacity to meet the demand for hospital beds and ICU beds (28.89 and 2.28 per 10,000, respectively). The additional use of moderate NPIs (20% home isolation and L1 PHSMs) could further reduce the number of deaths to levels lower than seasonal influenza (1.16 per 10,000), albeit still higher for 60+ individuals (4.67 deaths per 10,000). This combined strategy would also reduce the pressure on the health care system to manageable levels (20.19 hospital beds per 10,000 and 1.32 ICU beds per 10,000).

In the reference scenario, the 1.70 projected deaths per 10,000 individuals corresponds to a 76-84% decrease compared with the estimated number of deaths during the Omicron BA.5 wave (7.08-10.62 per 10,000 [43]). This decrease is due to the increased protection caused by the BA.5 wave. Given the relatively long-lasting protection against severe disease and death conferred by vaccination or infection [10] (as opposed to the short-lasting protection against infection [8]), later seeding of the epidemic would lead to a relatively small increase in the death toll (Supplementary Figure 2). The estimated decrease in the incidence of deaths between two successive BA.5 and XBB waves is consistent with the 55% reduction reported by the Singapore surveillance system [12]. In absolute terms, it is difficult to compare the incidence of deaths projected by our model for Shanghai with that observed in Singapore (0.17 deaths per 10,000 individuals), as the landscape immunity of the two populations is very different. For instance, different vaccine products were used to immunize the two populations, and the history of previous infections (with China maintaining a “zero-COVID” policy until late 2022), population demographics, and contact patterns also differed. Moreover, during the XBB wave, Singapore relied heavily on the distribution of antiviral drugs and NPIs.

To accurately interpret our findings, it is important to consider that our study focuses on Shanghai. Due to variations in factors such as demographic characteristics, contact patterns, vaccination rates, and natural immunity across different regions of China, it would be difficult to draw definitive conclusions about the entire country based on our analysis. However, Shanghai had one of the lowest vaccination coverages in China during the COVID-19 pandemic in December 2022 [44], which makes it a useful worst-case scenario for assessing the potential impact of XBB.1.5.

Furthermore, our study has several limitations that should be considered. First, the clinical severity of XBB.1.5 is still not fully understood, and we assumed that it would be similar to that of BA.5. As a result, we may have overestimated the burden of XBB.1.5. Second, the protection and duration of immunity against different clinical endpoints for Omicron XBB.1.5 is not yet fully known. To account for this uncertainty, we conducted sensitivity analyses to explore the variant’s potential impact. Finally, while we made educated assumptions and conducted multiple sensitivity analyses on the net reproduction number of a possible XBB.1.5 wave based on its growth rate in other locations and the growth rate of previous Omicron subvariants in China, it is possible that a future XBB.1.5 wave could spread at a different rate.

In conclusion, our study shows that a potential resurgence of COVID-19 caused by Omicron XBB.1.5 in Shanghai is less likely to overwhelm the health care system and would result in a death toll comparable to that of seasonal influenza. To achieve these outcomes, however, maintaining a combination of vaccination, antiviral treatments, and moderate levels of NPIs is needed. Hence, the development and use of highly effective vaccines with long-term immune persistence and antiviral treatments remain key public health priorities to further reduce disease burden and pressure on the health care system and to lower the reliance on NPIs.

## Data Availability

The code and data used to conduct these analyses are found at https://github.com/HengcongLiu/Shanghai-XBB.

## Declarations

### Authors’ contributions

H.Y. conceived and designed the study. H.Y., and M.A. supervised the study. H.L. designed and developed the model. H.L. analyzed the model outputs. H.L. prepared the first draft of the manuscript. H.L., X.X., X.D., and L.Y. prepared the tables and figures. H.L., Z.H., R.S., JY.Z., J.D., Q.W., and X. C. participated in data collection and updated the relative literatures. H.Y., M.A., J.C., and JJ. Z. revised the content critically. All authors contributed to review and revision and approved the final manuscript as submitted and agree to be accountable for all aspects of the work.

### Ethics approval and consent to participate

Not applicable.

## Acknowledgements

Not applicable.

## Consent for publication

Not applicable.

