## Supplemental Table 1-4, Supplemental Figure 1-7 for "Projecting the potential impact of an Omicron XBB.1.5 wave in Shanghai, China"

### Method

We developed an age-structured compartmental model to simulate the transmission of Omicron XBB.1.5 in Shanghai, which is described by the following system of differential equations:

$$\frac{dS_{a}(t)}{dt}=-\left( 1-\varepsilon_{S} \right)S_{a}(t)\lambda_{a}(t)$$

$$\frac{dH_{a}(t)}{dt}=-\left( 1-\varepsilon_{H} \right)H_{a}\left( t \right)\lambda_{a}\left( t \right)-\omega_{1}H_{a}\left( t \right)$$

$$\frac{dW_{a}(t)}{dt}=\omega_{1}H_{a}\left( t \right)-\left( 1-\varepsilon_{W} \right)W_{a}\left( t \right)\lambda_{a}\left( t \right)-\alpha_{a}W_{a}\left( t \right)$$

$$\frac{dB_{a}(t)}{dt}=\alpha_{a}W_{a}\left( t \right)-\left( 1-\varepsilon_{B} \right)B_{a}\left( t \right)\lambda_{a}\left( t \right)-\omega_{2}B_{a}\left( t \right)$$

$$\frac{dO_{a}(t)}{dt}=\omega_{2}B_{a}\left( t \right)-(1-\varepsilon_{O})O_{a}(t)\lambda_{a}(t)$$

$$\frac{dE_{a}(t)}{dt}=\sum_{X\in\{S,H,W,B,O\}} \left( 1-\varepsilon_{X} \right)X_{a}\left( t \right)\lambda_{a}\left( t \right)-\gamma_{E}E_{a}(t)$$

$$\frac{dI_{a}(t)}{dt}=\gamma_{E}E_{a}\left( t \right)-\gamma_{R}I_{a}\left( t \right)$$

$$\frac{dR_{a}(t)}{dt}=\gamma_{R}I_{a}\left( t \right)$$

where:

- $S_{a}(t)$, $E_{a}(t)$, $I_{a}(t)$ and $R_{a}(t)$ represent the number of susceptible, latent, infectious and removed individuals in age group $a$ at time $t$.
- $H_{a}(t)$, $W_{a}(t)$, $B_{a}(t)$ and $O_{a}(t)$ represent the number of susceptible individuals with hybrid immunity, waned hybrid immunity, boosted hybrid immunity, and waned boosted hybrid immunity in age group $a$ at time $t$.
- $\varepsilon_{X}$ is the vaccine effectiveness against infection for individuals in state $X$.
- $\lambda_{a}(t)$ is the force of infection for age group $a$ at time $t$.
- $1/\gamma_{E}$ and $1/\gamma_{R}$ represent the latent and infectious period; since they are both exponentially distributed, their sum corresponds to the generation time [1].
- $1/\omega_{1}$ and $1/\omega_{2}$ are the interval between the latest infection and administration of a booster and the interval between the administration of a booster and waned boosted immunity, respectively.
- $\alpha_{a}$ is the rate at which age group $a$ receives a booster.

The time- and age-dependent force of infection $\lambda_{a}(t)$ is defined as:

$$\lambda_{a}\left( t \right)=\beta_{0}(1-\sigma)\sum_{b=1}^{m} C_{ab}\frac{\left( 1-r\varphi\right)I_{b}\left( t \right)}{N_{b}}$$

where:

- $\beta_{0}$ is the transmission risk, which determines the pathogen transmissibility in the absence of nonpharmaceutical interventions and vaccination.
- $\sigma$ represents the reduction in transmissibility ascribable to Level 1 PHSMs.
- $\varphi$ represents the fraction of infected individuals who are isolated at home.
- $r$ represents the reduction in transmissibility for infectious individuals isolated at home.
- $C_{ab}$ is the age-group-specific contact matrix, whose elements describe the mean daily numbers of contacts that a person in age group $a$ has with individuals in age group $b$.
- $N_{a}$ is the number of individuals in age group $a$.
- $m$ is the number of age groups.

The infection incidence of individuals in stats $X$, age group $a$ at time $t$ was calculated as follows:

$$A_{X,a,i}\left( t \right)=\left( 1-\varepsilon_{X} \right)X_{a}\left( t \right)\lambda_{a}\left( t \right)$$

Similarly, hospital admissions, ICUs, and deaths were calculated as follows:

$$A_{X,a,h}\left( t \right)=p_{a,h}\int_{0}^{t} \left( 1-\varepsilon_{X} \right)X_{a}\left( x \right)\lambda_{a}\left( x \right)(1-\theta_{a}\phi_{h})(1-\varepsilon_{X}^{H})f_{h}(t-x)dx$$

$$A_{X,a,u}\left( t \right)=p_{a,u}\int_{0}^{t} \left( 1-\varepsilon_{X} \right)X_{a}\left( x \right)\lambda_{a}\left( x \right)(1-\theta_{a}\phi_{u})(1-\varepsilon_{X}^{U})f_{u}(t-x)dx$$

$$A_{X,a,d}\left( t \right)=p_{a,d}\int_{0}^{t} \left( 1-\varepsilon_{X} \right)X_{a}\left( x \right)\lambda_{a}\left( x \right)(1-\theta_{a}\phi_{d})(1-\varepsilon_{X}^{D})f_{d}(t-x)dx$$

where:

- $p_{a,h}$, $p_{a,u}$ and $p_{a,d}$ are the infection hospitalization risk, infection ICU risk, and infection fatality risk for unvaccinated individuals in age group $a$, respectively.
- $\theta_{a}$ is the antiviral coverage for age group $a$.
- $\phi_{h}$, $\phi_{u}$ and $\phi_{d}$ are the antivirals effectiveness in reducing the risk of hospitalization, ICU, and death, respectively.
- $f_{h}$, $f_{u}$ and $f_{d}$ are the probability density functions of the time interval between infection and hospitalization, ICU admission, and death, respectively. $f_{h}$ and $f_{u}$follow a gamma distribution with a mean of 6.7 days and a standard deviation of 2.6 days; $f_{d}$ follows a gamma distribution with a mean of 22.3 days and a standard deviation of 9.5 days [2-4].
- $\varepsilon_{X}^{H}$, $\varepsilon_{X}^{U}$ and $\varepsilon_{X}^{D}$ are the conditional vaccine effectiveness against hospitalization, ICU and death given infection for individuals in state $X$, respectively.

### Supplementary Table 1: Protection of hybrid immunity against different clinical endpoints.

| Scenario | Clinical endpoint | Protection of hybrid immunity against different clinical endpoints *t* days after recovered from previous infection or after receiving a booster dose | | | |
| --- | --- | --- | --- | --- | --- |
|  |  | *t*=0 | *t*=90 | *t*=180 | *t*=360 |
| Hybrid immunity  (short duration) | Infection | 80% ^a^ | 60% | 40% ^[5]^ | 20% |
|  | Hospital admission or severe disease | 95% | 95% | 95% | 95% |
| Hybrid immunity  (long duration) | Infection | 80% ^a^ | 67% | 56% | 40% ^[5]^ |
|  | Hospital admission or severe disease | 95% | 95% | 95% | 95% |
| Boosted with monovalent  (assumed) | Infection | 80% | 60% | 40% | 20% |
|  | Hospital admission or severe disease | 95% | 95% | 95% | 95% |
| Boosted with bivalent  (assumed) | Infection | 85% | 65% | 45% | 25% |
|  | Hospital admission or severe disease | 98% | 98% | 98% | 98% |

^a^ Following the widely used method developed by *Khoury et al* [6], we estimated the protection of hybrid immunity against Omicron XBB.1.5 infection to be around 80% at 4 weeks after hospital discharge [7].

### Supplementary Table 2: Clinical severity of Omicron XBB.1.5

| **Age group (years)** | **IHR** | **IUR ^a^** | **IFR** |
| --- | --- | --- | --- |
| 0-2 | 0.01% | 0.001% | 0.0005% |
| 3-17 | 0.02% | 0.001% | 0.0005% |
| 18-29 | 0.61% | 0.001% | 0.0005% |
| 30-39 | 2.34% | 0.038% | 0.0230% |
| 40-49 | 2.94% | 0.038% | 0.0230% |
| 50-59 | 5.52% | 0.210% | 0.1260% |
| 60-69 | 7.98% | 0.210% | 0.1260% |
| 70-79 | 11.28% | 4.878% | 2.0000% |
| 80+ | 12.48% | 11.905% | 8.7000% |
| total | 4.08% | 0.802% | 0.4670% |

^a^ IUR was estimated by scaling the IFR using the ratio of IUR and IFR reported by Ministry of Health in Singapore during XBB wave [8].

### Supplementary Table 3: Reduction in hospital admission, ICU admissions and deaths compared to the reference scenario when varying the intensity of each intervention separately.

| **Scenario** | **Hospital**  **admissions (%)** | | **ICU**  **Admissions (%)** | | **Deaths**  **(%)** | |
| --- | --- | --- | --- | --- | --- | --- |
|  | Total | 60+ years old | Total | 60+ years old | Total | 60+ years old |
| 30% coverage | 1.2 | 4.9 | 7.7 | 8.2 | 9.5 | 9.7 |
| 90% coverage | -0.6 | -3.5 | -4.6 | -5.3 | -8.5 | -8.7 |
| 0% antivirals | 2.6 | 2.6 | 2.8 | 2.7 | 9.6 | 9.4 |
| 20% antivirals | -2.6 | -2.6 | -2.8 | -2.8 | -9.6 | -9.3 |
| No L1 PHSMs | 7.9 | 15.9 | 22.9 | 24.7 | 29.4 | 30.3 |
| 0% isolation | 3.6 | 7.1 | 10.0 | 10.8 | 13.0 | 13.3 |
| 20% isolation | -4.0 | -7.7 | -10.3 | -11.1 | -14.0 | -14.3 |

### Supplementary Table 4: Projected healthcare demand and number of deaths when combined interventions are adopted.

| **Vaccine**  **coverage** | **Antiviral**  **drugs** | **Isolation** | **Level 1**  **PHSMs** | **Peak prevalence**  **of hospital admissions**  **per 10,000** | **Peak prevalence**  **of ICUs**  **per 10,000** | **Cumulative incidence**  **of deaths per 10,000** | |
| --- | --- | --- | --- | --- | --- | --- | --- |
|  |  |  |  |  |  | Total | 60+ years old |
| 30% | 0% | 0% | No | 33.01 | 2.88 | 2.89 | 11.64 |
| 30% | 0% | 0% | Yes | 28.20 | 2.19 | 2.28 | 9.16 |
| 30% | 0% | 10% | No | 30.82 | 2.56 | 2.62 | 10.53 |
| 30% | 0% | 10% | Yes | 25.98 | 1.93 | 2.02 | 8.07 |
| 30% | 0% | 20% | No | 28.54 | 2.24 | 2.32 | 9.32 |
| 30% | 0% | 20% | Yes | 23.67 | 1.68 | 1.76 | 7.07 |
| 30% | 10% | 0% | No | 32.21 | 2.82 | 2.65 | 10.71 |
| 30% | 10% | 0% | Yes | 27.50 | 2.14 | 2.09 | 8.38 |
| 30% | 10% | 10% | No | 30.06 | 2.50 | 2.40 | 9.66 |
| 30% | 10% | 10% | Yes | 25.34 | 1.88 | 1.84 | 7.38 |
| 30% | 10% | 20% | No | 27.83 | 2.19 | 2.12 | 8.53 |
| 30% | 10% | 20% | Yes | 23.07 | 1.64 | 1.60 | 6.45 |
| 30% | 20% | 0% | No | 31.40 | 2.75 | 2.41 | 9.74 |
| 30% | 20% | 0% | Yes | 26.81 | 2.08 | 1.89 | 7.60 |
| 30% | 20% | 10% | No | 29.30 | 2.43 | 2.18 | 8.79 |
| 30% | 20% | 10% | Yes | 24.69 | 1.83 | 1.67 | 6.71 |
| 30% | 20% | 20% | No | 27.15 | 2.13 | 1.92 | 7.74 |
| 30% | 20% | 20% | Yes | 22.50 | 1.60 | 1.45 | 5.83 |
| 70% | 0% | 0% | No | 31.74 | 2.61 | 2.65 | 10.66 |
| 70% | 0% | 0% | Yes | 26.95 | 1.99 | 2.09 | 8.38 |
| 70% | 0% | 10% | No | 29.54 | 2.32 | 2.38 | 9.58 |
| 70% | 0% | 10% | Yes | 24.77 | 1.74 | 1.86 | 7.41 |
| 70% | 0% | 20% | No | 27.24 | 2.04 | 2.12 | 8.50 |
| 70% | 0% | 20% | Yes | 22.42 | 1.50 | 1.58 | 6.35 |
| 70% | 10% | 0% | No | 30.97 | 2.55 | 2.44 | 9.83 |
| 70% | 10% | 0% | Yes | 26.27 | 1.93 | 1.91 | 7.67 |
| 70% | 10% | 10% | No | 28.80 | 2.26 | 2.19 | 8.79 |
| 70% | 10% | 10% | Yes | 24.15 | 1.71 | 1.70 | 6.79 |
| 70% | 10% | 20% | No | 26.59 | 1.98 | 1.94 | 7.79 |
| 70% | 10% | 20% | Yes | 21.86 | 1.46 | 1.44 | 5.79 |
| 70% | 20% | 0% | No | 30.21 | 2.48 | 2.22 | 8.97 |
| 70% | 20% | 0% | Yes | 25.62 | 1.89 | 1.74 | 6.98 |
| 70% | 20% | 10% | No | 28.09 | 2.21 | 2.00 | 8.04 |
| 70% | 20% | 10% | Yes | 23.56 | 1.65 | 1.52 | 6.13 |
| 70% | 20% | 20% | No | 25.92 | 1.93 | 1.77 | 7.09 |
| 70% | 20% | 20% | Yes | 21.32 | 1.43 | 1.30 | 5.23 |
| 90% | 0% | 0% | No | 30.37 | 2.41 | 2.49 | 10.01 |
| 90% | 0% | 0% | Yes | 25.55 | 1.84 | 1.93 | 7.73 |
| 90% | 0% | 10% | No | 28.26 | 2.15 | 2.24 | 9.00 |
| 90% | 0% | 10% | Yes | 23.45 | 1.61 | 1.69 | 6.75 |
| 90% | 0% | 20% | No | 25.93 | 1.88 | 1.97 | 7.90 |
| 90% | 0% | 20% | Yes | 21.24 | 1.40 | 1.44 | 5.72 |
| 90% | 10% | 0% | No | 29.64 | 2.35 | 2.28 | 9.16 |
| 90% | 10% | 0% | Yes | 24.92 | 1.80 | 1.76 | 7.07 |
| 90% | 10% | 10% | No | 27.55 | 2.09 | 2.06 | 8.27 |
| 90% | 10% | 10% | Yes | 22.88 | 1.57 | 1.53 | 6.15 |
| 90% | 10% | 20% | No | 25.30 | 1.84 | 1.80 | 7.24 |
| 90% | 10% | 20% | Yes | 20.72 | 1.36 | 1.30 | 5.18 |
| 90% | 20% | 0% | No | 28.89 | 2.28 | 2.08 | 8.37 |
| 90% | 20% | 0% | Yes | 24.28 | 1.74 | 1.60 | 6.45 |
| 90% | 20% | 10% | No | 26.87 | 2.03 | 1.87 | 7.53 |
| 90% | 20% | 10% | Yes | 22.30 | 1.53 | 1.39 | 5.56 |
| 90% | 20% | 20% | No | 24.66 | 1.80 | 1.63 | 6.57 |
| 90% | 20% | 20% | Yes | 20.19 | 1.32 | 1.16 | 4.67 |

### Supplementary Figure 1: Intrinsic generation time.

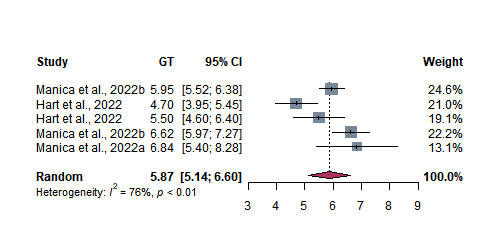

**Figure 1.** Pooled estimates of the intrinsic generation time based on references [9-11].

### Supplementary Figure 2: Sensitivity analysis on the seeding date

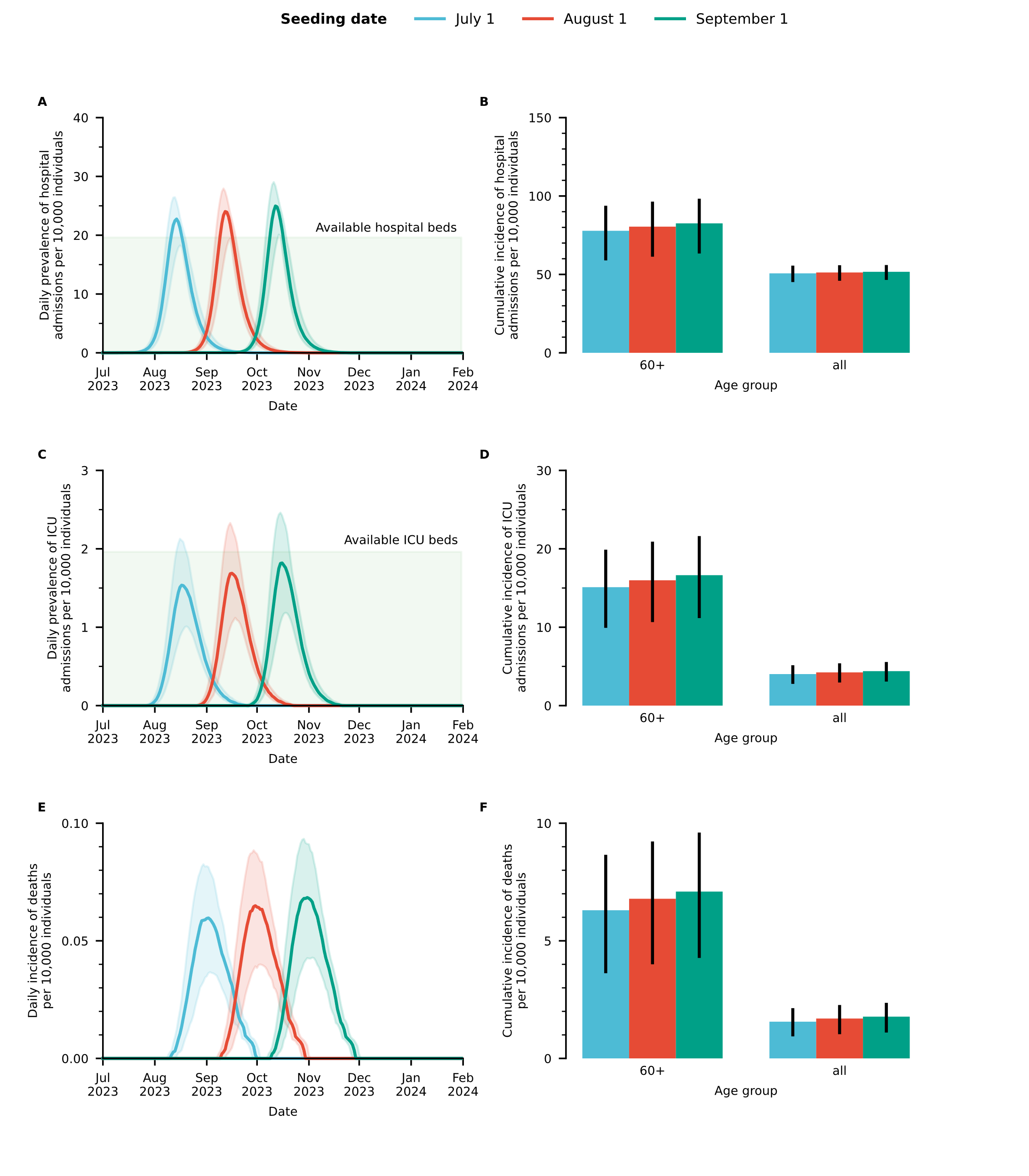

**Figure 2**. A. Daily prevalence of hospital admissions per 10,000 individuals. The shaded area corresponds to the maximum hospital bed capacity. B. Cumulative number of hospital admissions by age group per 10,000 individuals in that age group. C. Daily prevalence of ICU admissions per 10,000 individuals. The shaded area corresponds to the maximum ICU capacity. D. Cumulative number of ICU admissions by age group per 10,000 individuals in that age group. E. Daily incidence of deaths per 10,000 individuals. F. Cumulative number of deaths by age group per 10,000 individuals in that age group. Data are presented as median and 95% CIs of 100 stochastic model realizations.

### Supplementary Figure 3: Sensitivity analysis on the number of imported infections

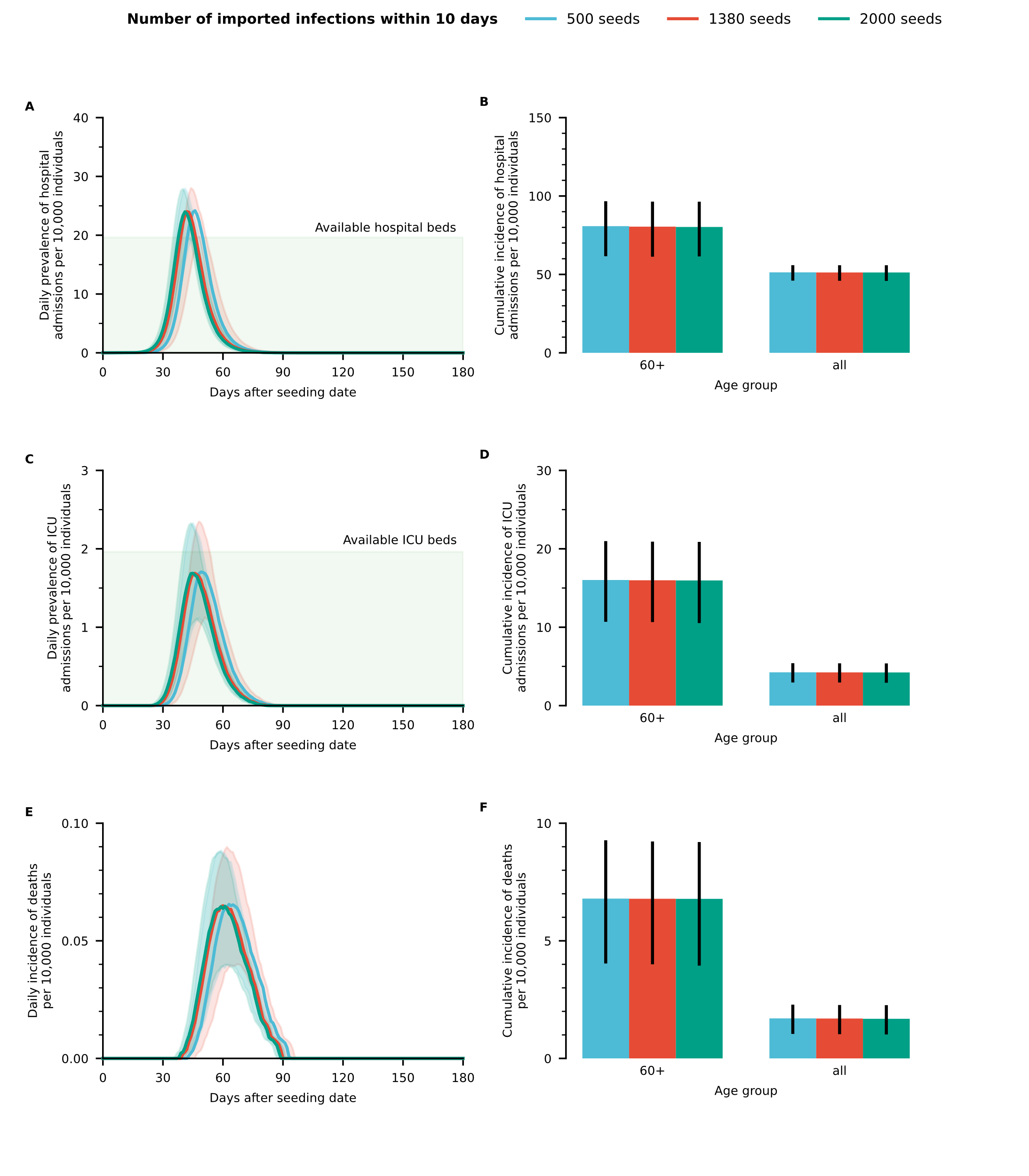

**Figure 3.** A. Daily prevalence of hospital admissions per 10,000 individuals. The shaded area corresponds to the maximum hospital bed capacity. B. Cumulative number of hospital admissions by age group per 10,000 individuals in that age group. C. Daily prevalence of ICU admissions per 10,000 individuals. The shaded area corresponds to the maximum ICU capacity. D. Cumulative number of ICU admissions by age group per 10,000 individuals in that age group. E. Daily incidence of deaths per 10,000 individuals. F. Cumulative number of deaths by age group per 10,000 individuals in that age group. Data are presented as median and 95% CIs of 100 stochastic model realizations.

### Supplementary Figure 4: Sensitivity analysis on the effective reproduction number

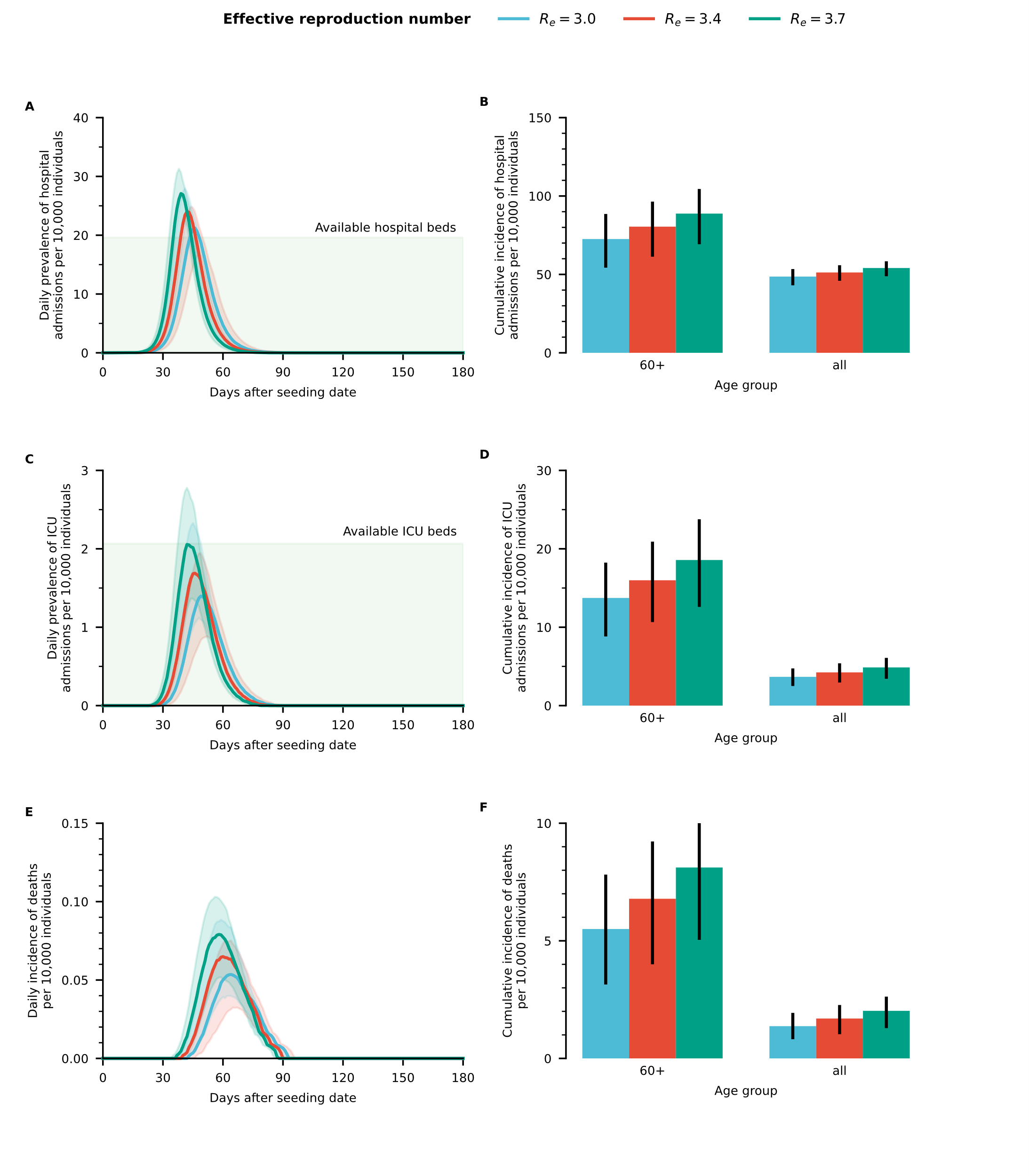

**Figure 4.** A. Daily prevalence of hospital admissions per 10,000 individuals. The shaded area corresponds to the maximum hospital bed capacity. B. Cumulative number of hospital admissions by age group per 10,000 individuals in that age group. C. Daily prevalence of ICU admissions per 10,000 individuals. The shaded area corresponds to the maximum ICU capacity. D. Cumulative number of ICU admissions by age group per 10,000 individuals in that age group. E. Daily incidence of deaths per 10,000 individuals. F. Cumulative number of deaths by age group per 10,000 individuals in that age group. Data are presented as median and 95% CIs of 100 stochastic model realizations.

### Supplementary Figure 5: Sensitivity analysis on waning rate of hybrid immunity

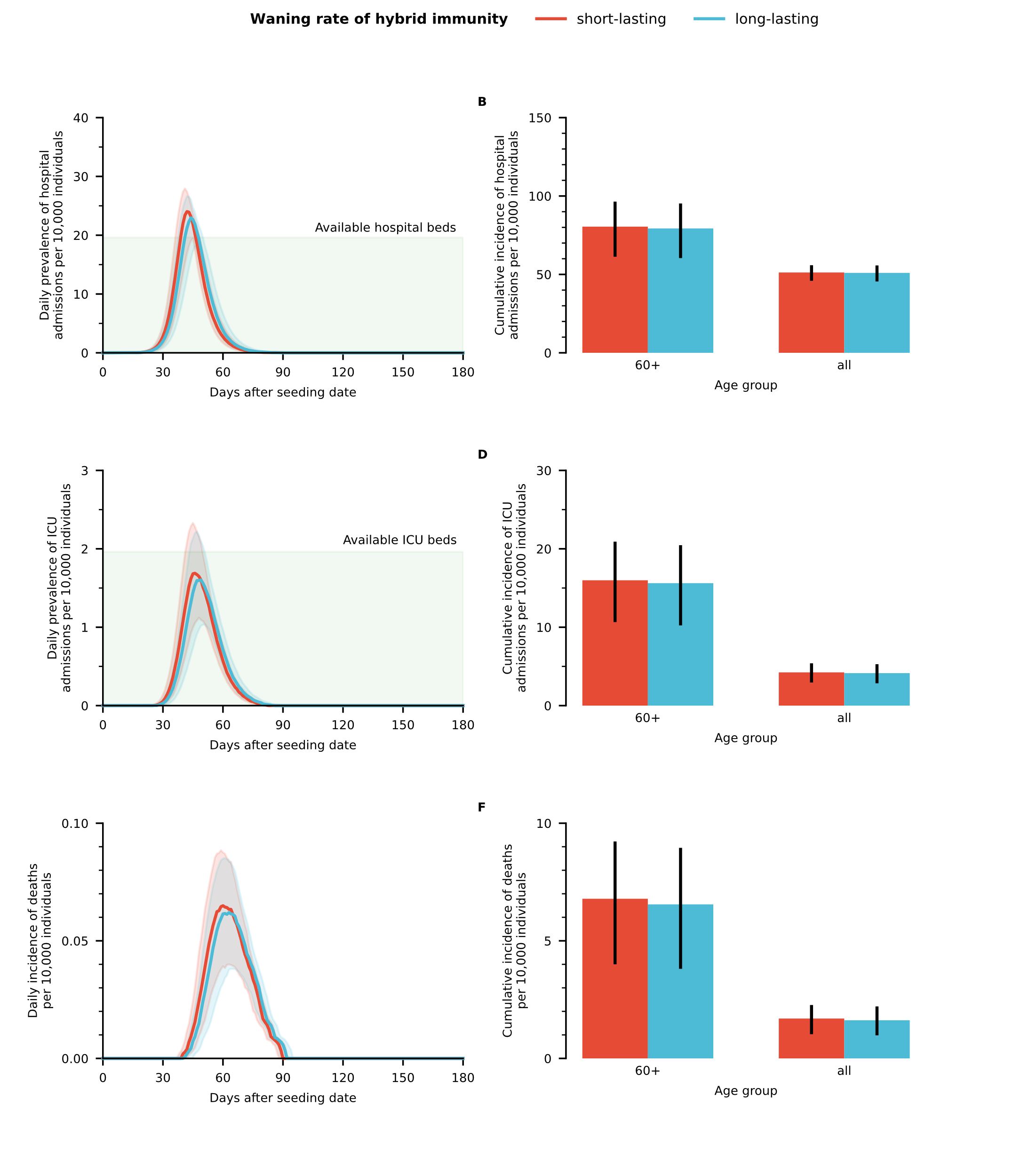

**Figure 5.** A. Daily prevalence of hospital admissions per 10,000 individuals. The shaded area corresponds to the maximum hospital bed capacity. B. Cumulative number of hospital admissions by age group per 10,000 individuals in that age group. C. Daily prevalence of ICU admissions per 10,000 individuals. The shaded area corresponds to the maximum ICU capacity. D. Cumulative number of ICU admissions by age group per 10,000 individuals in that age group. E. Daily incidence of deaths per 10,000 individuals. F. Cumulative number of deaths by age group per 10,000 individuals in that age group. Protection against infection was considered to last 6 and 12 months on average in the baseline and long-lasting scenario, respectively. Data are presented as median and 95% CIs of 100 stochastic model realizations.

### Supplementary Figure 6: Sensitivity analysis on booster vaccine

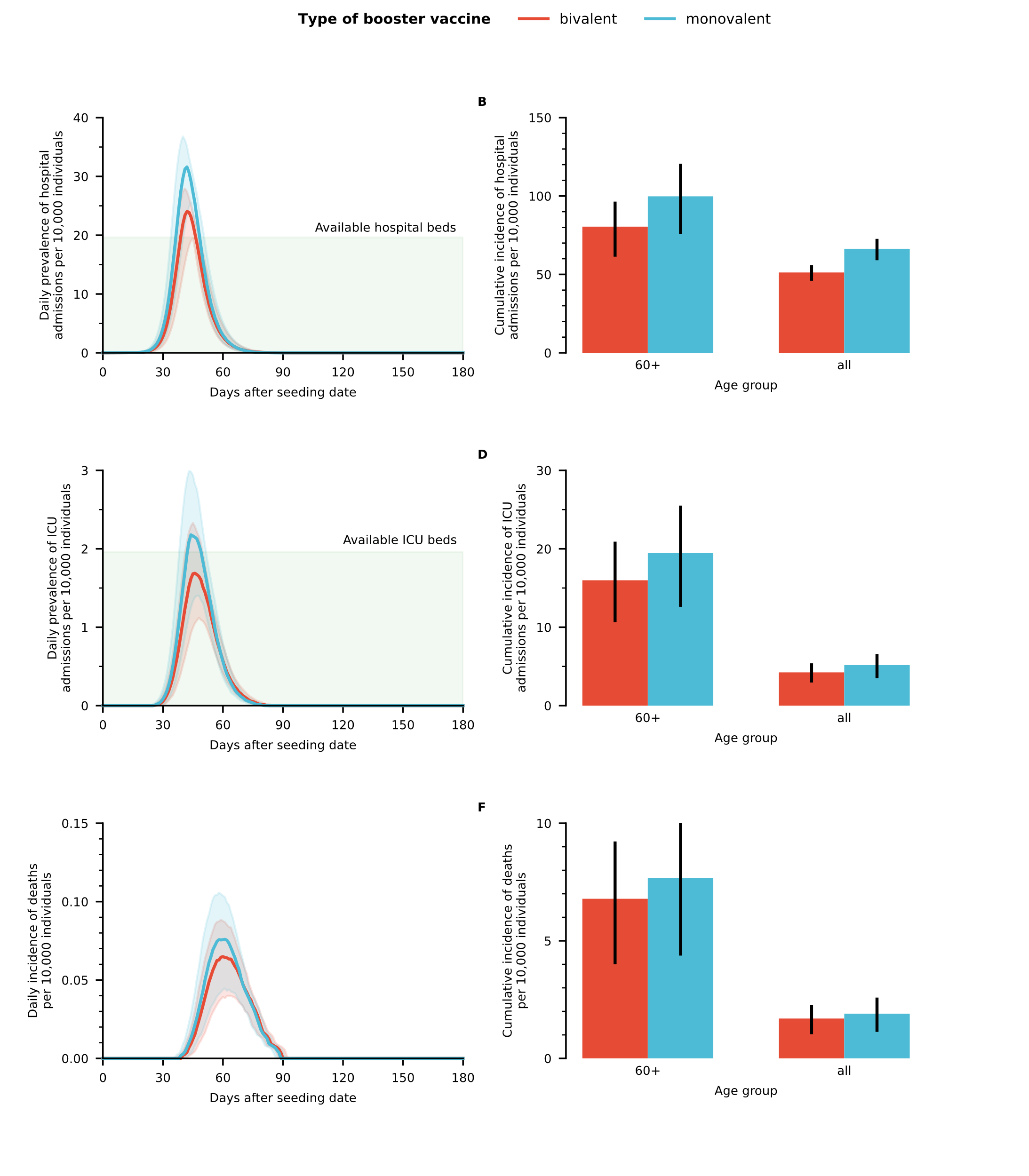

**Figure 6.** A. Daily prevalence of hospital admissions per 10,000 individuals. The shaded area corresponds to the maximum hospital bed capacity. B. Cumulative number of hospital admissions by age group per 10,000 individuals in that age group. C. Daily prevalence of ICU admissions per 10,000 individuals. The shaded area corresponds to the maximum ICU capacity. D. Cumulative number of ICU admissions by age group per 10,000 individuals in that age group. E. Daily incidence of deaths per 10,000 individuals. F. Cumulative number of deaths by age group per 10,000 individuals in that age group. Protection against infection was considered to last 6 and 12 months on average in the baseline and long-lasting scenario, respectively. Data are presented as median and 95% CIs of 100 stochastic model realizations.

### Supplementary Figure 7: Projected demand of hospital beds and ICU beds: varying the intensity of each intervention separately.

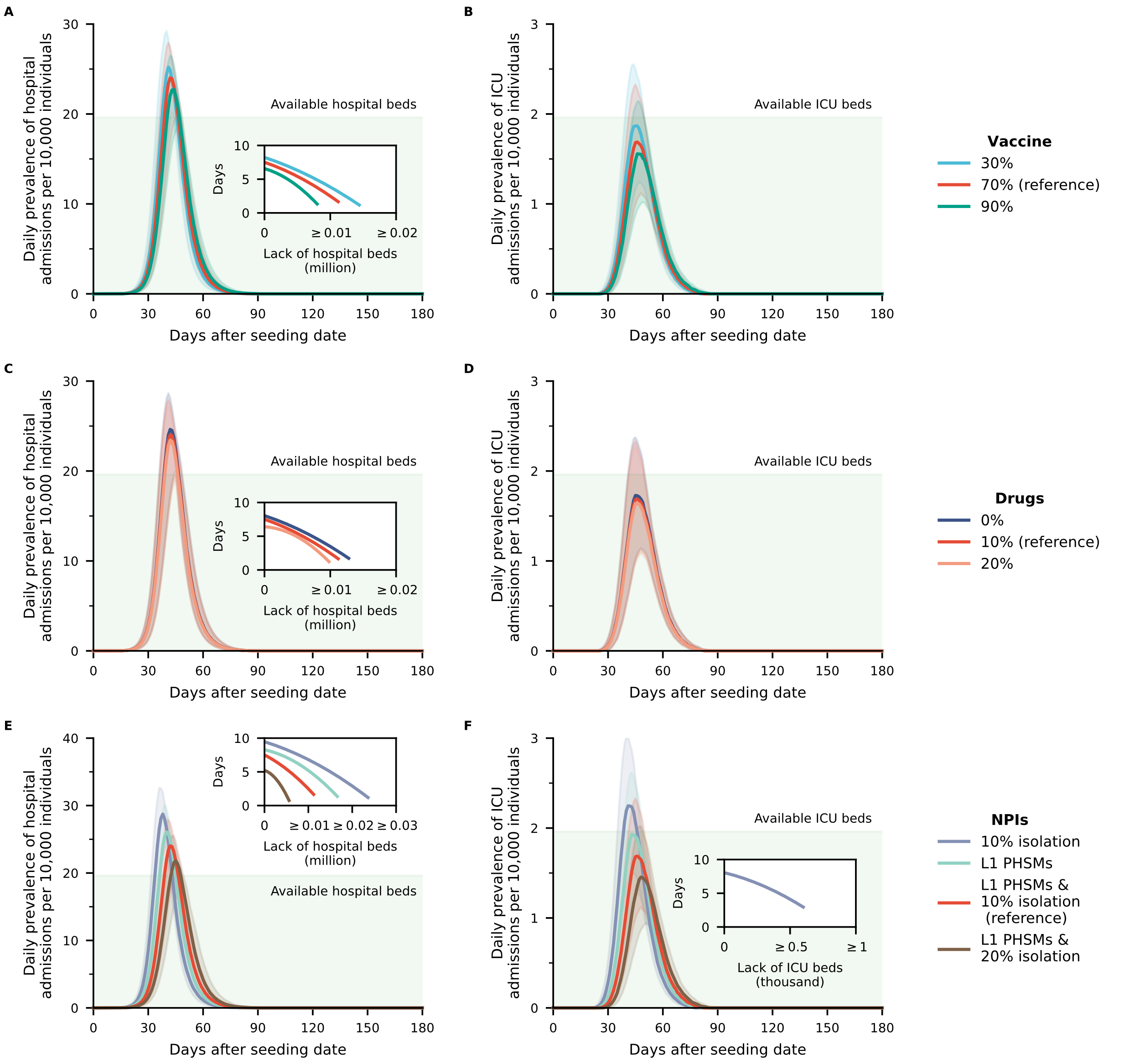

**Figure 7.** A Daily prevalence of hospital admissions per 10,000 individuals under different scenarios of bivalent booster coverage. The shaded area corresponds to the maximum hospital bed capacity. The inset shows the number of days of hospital beds shortage as a function of the number of missing beds. B Same as A, but for ICU. C-D Same as A-B, but for scenarios considering different coverages of antiviral drugs. E-F Same as A-B, but for scenarios exploring different NPIs. *Reference* represents the reference scenario considering 70% booster vaccine coverage, 10% antiviral drugs, 10% home isolation, and L1 PHSMs are implemented. The other scenarios are obtained by changing a single parameter at the time as compared to the reference scenario. Data are presented as median and 95% CIs of 100 stochastic model realizations.
